# Understanding the Bias between the Number of Confirmed Cases and Actual Number of Infections in the COVID-19 Pandemic

**DOI:** 10.1101/2020.06.22.20137208

**Authors:** Xingang Chen, Dhiraj Kumar Hazra

## Abstract

The number of positive cases confirmed in the viral tests is a probe of the actual number of infections of COVID-19. The bias between these two quantities is a key element underlying the determination of some important parameters of this disease and the policy-making during the pandemic. To study the dependence of this bias on measured variables, we introduce a parameterization model that motivates a method of organizing the daily data of the numbers of the total tests, confirmed cases, hospitalizations and fatalities. After comparing with the historical data of the USA in the past few months, we find a simple formula relating these four variables. As a few applications, we show, among other things, how this formula can be used to project the number of actual infections, to provide guidance on how the test volume should be adjusted, and to derive an upper bound on the overall infection fatality rate of COVID-19 (*<* 0.64%, 95% C.L.) and a theoretical estimate of its value.

## 1 Introduction

It is difficult to estimate the total number people infected with COVID-19. What each region or country directly measures is the number of infections among those who are tested. For many reasons, the people who get tested include only part of the actual infected. For example, the total number of tests may be limited; among suspected infections, only those with more severe symptoms are recommended or have the motivation to take the tests. So, there is a bias between the number of confirmed cases and the actual number of infections. The bias presents a barrier to estimate the number of people that are infected but untested, therefore, is a key element to be understood in order to estimate some important parameters of the disease, such as the infection fatality rate, and in order to decide on the policies of testing strategies, lockdown and economy reopening. The bias even interferes with the function of the viral tests as a monitor of the daily development of the pandemic, because the number of confirmed cases not only depend on the actual number of infections, but also on the test volume.

In this paper, we try to understand more quantitatively the dependence of this bias on the several aforementioned variables. We introduce a simple model that relates these variables in section 2, and compare the model with the available data reported from different states of the USA in the past few months in section 3. These data include the historical data of the daily numbers of the total tests, confirmed cases, hospitalization, and fatalities. We find a simple formula relating these four variables. In section 4, we show several applications of our results, including how the model can be used to estimate the actual number of infections, to predict some important parameters of COVID-19, such as the infection fatality rate, based on limited information, and to provide guidance on how much the test volume should be expanded. In section 5, we discuss some issues related to the analyses and on how the method and results may be applied in a wider context and developed in future works.

## 2 Modelling the bias

### 2.1 Normalising number of hospitalizations

Before discussing the bias, we first introduce the roles of the number of hospitalizations and fatalities in this analyses.

Let us denote the total actual number of infections each day as *N* (*t*). (This function, and other functions we will introduce later, are functions of time *t* in terms of days. For simplicity, in most cases, we will not explicitly indicate the *t* variable in these functions.) Although *N* is a number that is difficult to measure directly, its value may be proportional to some variables with the proportional constants determined by the intrinsic properties of the disease and so independent of the viral testing. There are two choices of such tracers of *N*: the daily number of new hospitalizations and new fatalities. Both are good candidates, as long as the resource of hospitals is not stretched, so that all patients who are severely ill can be hospitalized in a timely manner. To be a good tracer, we would like to have the conditions leading to hospitalization or fatality as uniform as possible across different hospitals and regions.

To examine the uniformity of these conditions, we first notice that the fatality rates per unit hospitalization, namely the hospitalization fatality rates (HFRs), computed in Appendix A, are slightly different across the states in the USA. This means that at least one of the aforementioned conditions is slightly different across the states. Among these two choices, it may be more reasonable to assume that the condition leading to fatality is more likely to be uniform than that to hospitalization. Therefore, it would be a better choice to assume that the actual number of infections is proportional to the number of fatalities.

However, using the number of fatalities as the tracer has a disadvantage that, on average for the same patient, there is likely a significant time-delay between the report dates of the viral test and the fatality. On the other hand, such a delay, if any, would be much shorter in the case of hospitalization. To take the advantages of both possibilities, we normalize the values of *H* in such a way that, with the normalized values *H*_*N*_, all states have the same HFR. We then use *H*_*N*_ as the tracer and assume *N ∝ H*_*N*_. In this way, we have not only effectively used the better choice, i.e. the number of fatalities, as the tracer, but also significantly reduced or eliminated the time delay. This normalization procedure is described in Appendix A.

We note that, although this normalization procedure on *H* eliminates some ambiguities, we have checked that the main conclusion of our paper remain the same if we replace *H*_*N*_ with *H* in the following analyses. We will also proceed by first ignoring another possible time delay between the test result and hospitalization and discuss about this issue later.

### 2.2 Confusion factor in decisions for tests

The urge to take the viral test grows with the severeness of the COVID-19 symptoms. We introduce a variable *s* to label the severeness of the symptoms. The variable *s* runs from 0 to 1. If *s* is closer to 0, the symptoms are more distinct of COVID-19, such as loss of smell and taste, fatigue, persistent cough, and loss of appetite [1]. If *s* is closer to 1, the symptoms are more indistinguishable from common flu, such as mild cough and fever, or even without obvious symptoms (commonly known as the asymptomatic cases). When *s* is small, the patients are more likely to be recommended by doctors to take the tests, or to request to be tested. When *s* is large, the patients are more likely to stay home and rest without being tested. Such a variable may be generalized to multi-dimensional variables in a more sophisticated analysis.

The number of patients are distributed as a function of *s*, and the total number of infections is an integration from *s* = 0 to 1. To describe this distribution, we define a function *n*(*s*) to be the number of COVID-19 infections per unit *ds* per unit *H*_*N*_,

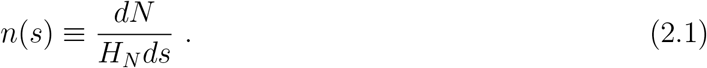

As discussed above, the symptoms of COVID-19 may be confused with those of common flu, and this confusion grows with *s*. We define a confusion factor *f* (*s*), which grows monotonically with *s* and reach a maximum number at *s* = 1. The maximum value *f* (1) may be close to infinity because asymptomatic cases are also included in *s* = 1, therefore, all population, including healthy people, are source of confusion. The number of people who get tested per unit *ds* is

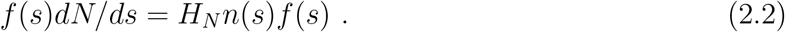

In realistic situations, not all these people between the entire range from *s* = 0 to 1 will be tested, and it is those with smaller *s* who have the priorities or motivations to get tested. This is the reason for the bias that we mentioned. We introduce a cutoff *s*_*c*_ such that only the people between *s* = 0 and *s* = *s*_*c*_ are tested.

### 2.3 Relations between various variables

With all the necessary functions introduced, we can now list the expressions for the following quantities.

- Total number of tests

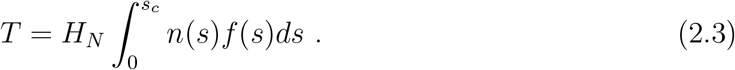
- Number of confirmed cases:

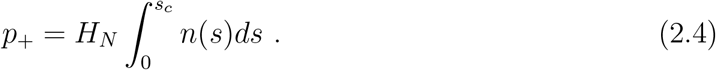
- Number of negative cases:

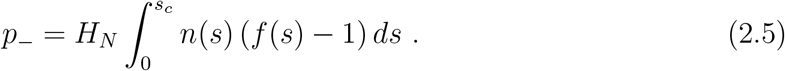
- Positive rate:

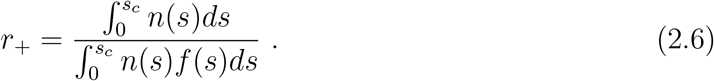
- Actual number of infections:

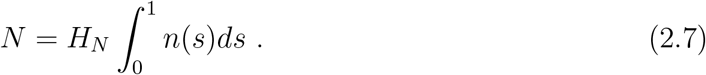

The expressions become simplified if we notice that we can always mathematically redefine the variable *s* such that *n*(*s*) = constant *≡ n*_0_, while keep *s* in the same [0, 1] range. We then have

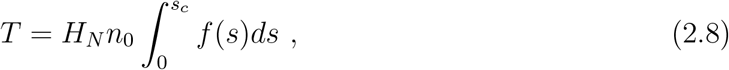

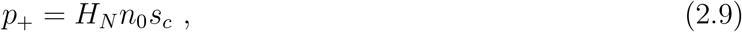

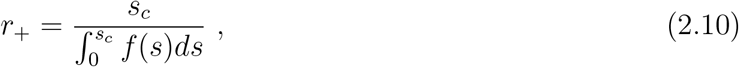

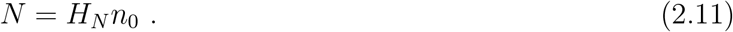

Using Eq. (2.8) and (2.9), we have

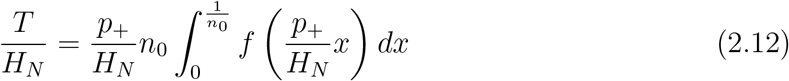

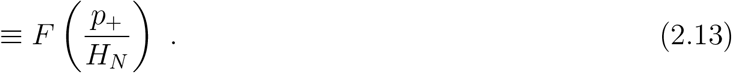

So, *T/H*_*N*_ can be expressed as a function of *p*_+_*/H*_*N*_ which contains some parameters. By fitting a template function with data, these parameters, and hence the function *F* (*p*_+_*/H*_*N*_), can be determined.

Before fitting with any data, there are some simple theoretical constraints on the behavior of this function. First, *T/H*_*N*_ = 0 when *p*_+_*/H*_*N*_ = 0. Second, because the daily actual number of infections is a very small fraction of the total population, *p*_+_*/H*_*N*_ should approach to a constant, which we denote as (*p*_+_*/H*_*N*_)_limit_, as *T/H*_*N*_ *→ ∞*. This constant equals to *N/H*_*N*_, where *N* is the definition of the actual number of infection in (2.11), because 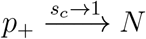 *N* and 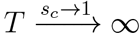.

The fact that we are interested in the quantities *T* and *p*_+_ divided by *H*_*N*_ can be readily understood as follows. Using *T* as an example, to capture more infected patients with less severe symptoms, increasing the total number of tests *T* is not a sufficient condition. One also has to consider the increase of the actual number of infections, which is assumed to be proportional to *H*_*N*_.

## 3 Data analyses

The four different daily variables important to our analyses are the number of total tests, confirmed cases, hospitalizations and fatalities. We use the data of the following states from the database of the Covid Tracking Project [2] (see also [3]), because these states provide all necessary information to extract these four quantities and at the same time have large values of daily new hospitalizations (*H ≫* 10) so that the statistical errors are relatively small: NY, FL, MA, GA, TN, OH, VA, MD, WI, MN, AZ.

A total of 805 state-days of data are collected. There are some non-positive numbers in the data that we discuss how to adjust in Appendix B. Besides that, we caution that there are still a small number of data that are likely affected by various artificial processes, for example, due to report delays and scheme changes. As long as these data are positive numbers, we include them as recorded in the database and we regard the associated errors as part of the statistical fluctuations and will discuss the treatment shortly.

The details on how Figure 1 is generated are described in Appendix C. A short summarize is as follows.

**Figure 1:**
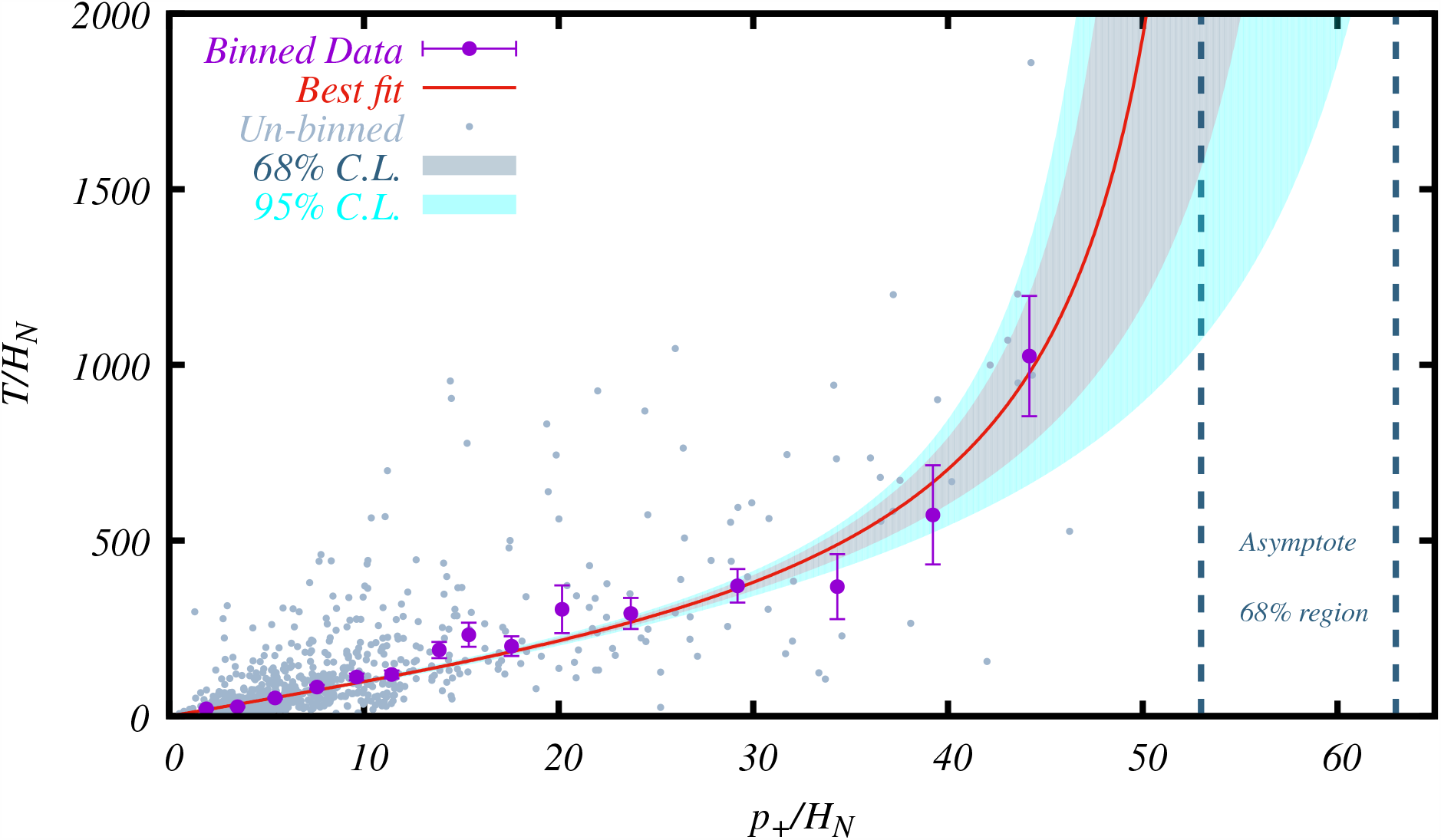
The relation between *T/H*_*N*_ and *p*_+_*/H*_*N*_ from 789 state-days of data from 11 USA states. Each of the 789 grey points represents the values computed from the data of a state-day. The purple bars indicate the mean values and the 1-sigma error-of-the-mean of the binned data points. The red line is the best-fit of the theoretical model (3.14), and the bands are its 1 and 2-sigma errors.

In order to analyze the relation between *T/H*_*N*_ and *p*_+_*/H*_*N*_, we generate the values of *p*_+_*/H*_*N*_ and *T/H*_*N*_ for each state-day and represent it as a dot using these values as the *x* and *y*-coordinate, respectively. This procedure gives rise to 805 points. The daily points associated with large positive rates are more likely to explore the lower-left part of the parameter space in this plot, while those with small positive rates are more likely to explore the upper-right part of the parameter space. The advantage to plot the distribution of the daily data, instead of studying just the average values of the states or the country, is that they not only provide the information about the average values, but also about daily variations that explore a much wider part of the parameter space, which helps us better understand the functional form of Eq. (2.13).

To find the form of Eq. (2.13), we bin the data and compute the mean value and the error of the mean for each bin. The mean values and their 1-sigma error-of-the-mean are shown as bars in the plots. After binning, the pattern of the data becomes much more clear. The binned data in *p*_+_*/H*_*N*_ *<* 50 follows the theoretical constraints stated below Eq. (2.13). The four binned data points (coming from 16 unbinned data point) in the *p*_+_*/H*_*N*_ *>* 50 region are much more scattered (see Figure 5 in Appendix C). Since large systematic errors are expected as mentioned previously, we discard these points that do not follow the theoretical constraint at large *T/H*_*N*_, and arrive at the final plot in Figure 1.

The binned data exhibits a trend that matches the theoretical constraints stated below Eq. (2.13), as well as a linear behavior near the origin. The simplest function that matches these behaviors are the tangent or inverse hyperbolic tangent function. We present the former case below, and leave the latter for Appendix C where more details of the data fitting procedures are described. For the tangent model,

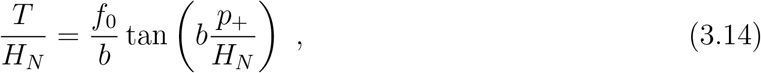

the best-fit parameters are *f*_0_ = 9.59, *b* = 0.0279. The 68% and 95% constaints and the correlation between the parameter *f*_0_ and *b* can be found in Figure 6 in Appendix C. For *bp*_+_*/H*_*N*_ *<* 1, Eq. (3.14) can be approximated by a polynomial series expansion,

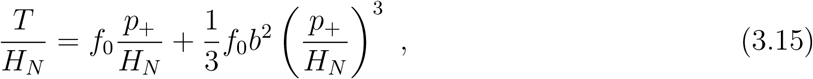

and this is sufficient to give a reasonable fit to all the data points. The analytical completion of this polynomial function with the tangent function (3.14), or the inverse hyperbolic tangent function, has the theoretical expectations built in, and the part of the functions beyond the current data range should be regarded as the theoretical prediction of the models. We therefore present two types of predictions of our results: one directly from data-fitting and another from theoretical extrapolation.

The last binned data point has *T/H*_*N*_ *≈* 1025, if we assume that the fitting function is only valid up to this point, from the error bands we can infer a lower bound for the asymptotic limit of *p*_+_*/H*_*N*_, ^1^

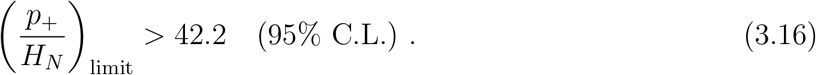

If we take the theoretical extrapolation of this model, we can get the asymptotic limit of *p*_+_*/H*_*N*_,

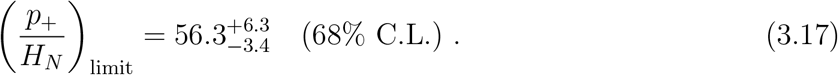

## 4 Applications

Once the function *T/H*_*N*_ = *F* (*p*_+_*/H*_*N*_) is found, the four variables, *T, p*_+_, *H* and *D*, are related in one formula, and there are many applications. We list a few examples in this section.

### 4.1 Estimating number of infections per hospitalization from viral tests

Viral tests can only detect a portion of all infections. Our results provide a way to quantitatively estimate this bias as soon as we know the positive rate of the test.

Given a positive rate, defined as

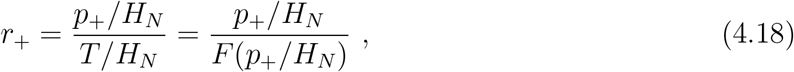

together with the obtained functional form of *F* (*p*_+_*/H*_*N*_), we can inversely solve for *p*_+_*/H*_*N*_, namely, the number of confirmed cases per normalized-hospitalization. Note that, due to the periodicity of tangent function, this is a multi-root problem and the relevant solution is the smallest positive root associated with the branch that is continuously connected to the point *p*_+_*/H*_*N*_ = *T/H*_*N*_ = 0.^2^ To compute the bias, we compare the ratio of *p*_+_*/H*_*N*_ at a given *r*_+_, which is the detected value of *p*_+_*/H*_*N*_, to that at *r*_+_ = 0, which equals to the value of *N/H*_*N*_. We can also compute the ratio between *p*_+_*/H*_*N*_ ‘s with different positive rates to get the bias between them. The results are presented in Table 1.

**Table 1:** The percentage of the actual number of infections detected in a test (i.e. the bias), given the positive rate of the test. The numbers in black are estimated from the results from the data fitting. The numbers in blue are based on the theoretical extrapolation Eq. (3.14). In both cases, best-fit parameters are used. The second row lists the number of confirmed patients per normalized-hospitalization, *p*_+_*/H*_*N*_, detected in each test with the corresponding positive rate. In the third row, we compute the bias of each test with respect to the test with *r*_+_ = 4%. We take 4% as the reference point because this is the best we can infer directly from the current data, without any theoretical extrapolation. In the fourth row, we compute the bias with respective to the actual number of infections. The fifth row lists the total number of tests per normalized-hospitalization, *T/H*_*N*_, performed in each test with the corresponding positive rate. In the sixth row, we compute how many tests are needed to achieve the goal of *r*_+_ = 2%, assuming the same actual number of infections. Note that a test with *r*_+_ = 2% is able to detect approximately 91% of the actual number of infections.

| positive rate ($r_+$) | 10% | 8% | 6% | 5% | 4% | 3% | 2% | 1% | $\sim 0\%$ |
| --- | --- | --- | --- | --- | --- | --- | --- | --- | --- |
| detected $p_+/H_N$ | 12.5 | 29.3 | 38.8 | 42.5 | 45.9 | 48.8 | 51.5 | 54.0 | 56.3 |
| bias (w.r.t. $r_+ = 4\%$) | 27% | 64% | 85% | 93% | 1 | 1.06 | 1.12 | 1.18 | 1.23 |
| bias (w.r.t. $r_+ = 0\%$) | 22% | 52% | 69% | 75% | 82% | 87% | 91% | 96% | 1 |
| $T/H_N$ performed | 125 | 366 | 646 | 851 | 1147 | 1628 | 2577 | 5402 | ∞ |
| tests needed to achieve $r_+ = 2\%$ | $\times 21$ | $\times 7.0$ | $\times 4.0$ | $\times 3.0$ | $\times 2.2$ | $\times 1.6$ | $\times 1$ | $\times 0.48$ | $\sim 0$ |

Once *p*_+_*/H*_*N*_ is known, *T/H*_*N*_ can be derived. Comparing the values of *T/H*_*N*_ at different *r*_+_ predicts how much the test volume needs to be increased to achieve a smaller *r*_+_ with the same actual number of infections. The results are also presented in Table 1.

Table 1 includes examples that are based on the results from the data fitting (numbers in black), as well as on the theoretical extrapolation from Eq. (3.14) beyond the data range (numbers in blue). As an example of how to interpret the numbers in this table, let us suppose that a daily test has been performed which yields a positive rate 10%. According to Table 1, such a test detects about 12.5 infections per normalized-hospitalization, which is only about 27% of the infections that could have been detected by a more aggressive test with a positive rate 4%, and about 22% of the actual number of infections. In addition, in order to achieve the positive rate *r*_+_ = 2% with the same actual number of infections, the test volume needs to be increased by a factor of 21. If we only would like to reduce the positive rate to *r*_+_ = 4%, then the test volume needs to be increased by a factor of 21*/*2.2 = 9.5.

Note that, according to the best-fit model, the highest positive rate accommodated is 1*/f*_0_ *≈* 10%. This means that, even the most distinct cases of COVID-19 infections have a confusion factor of about 10. This result reflects the USA’s national average situation, but may not be applied to some individual states. As we can see from Figure 8, states such as MA and NY have positive rates much higher than 10% at the small *p*_+_*/H*_*N*_ region, but these are balanced by the data from other states such as FL and WI, where the positive rates are much lower around the same parameter space.

### 4.2 Estimating infection hospitalization rate (IHR) and infection fatality rate (IFR)

The model predicts the actual number of infections per *H*_*N*_, which is given by the asymptotic limit of *p*_+_*/H*_*N*_. This value is important for estimating the values of the IHR and IFR of COVID-19. Again, we provide two kinds of estimates: one conservative estimate based on the data fitting, which is only able to give upper bounds on the values of IHR and IFR; and another based on the analytical extrapolation which can estimate the values of IHR and IFR.

The value of IHR depends on the standards for hospitalization, which may vary from state to state. In our convention, *H*_*N*_ is the hospitalization number normalized to the NY state standard assuming the conditions that lead to fatality is uniform for all states. The value of IHR for NY is *H*_*N*_ */N*, which is the inverse of (*p*_+_*/H*_*N*_)_limit_. Therefore, from the bound (3.16), we get the upper bound on the value of IHR,

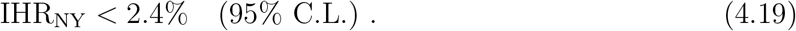

This value is based on the error bands at the last binned data point in Fig. 1. The value of *H*, as well as the value of IHR for other states, can be converted from *H*_*N*_ using the normalization factor *α* listed in Table 4, *H* = *H*_*N*_ */α* and so IHR = IHR_NY_*/α*.

To give an estimate on the actual value of IHR, we have to use a theoretical model that can be extrapolated to *T/H*_*N*_ *→ ∞*. From the model (3.14) and the asymptotic limit (3.17), we get the following mean value:

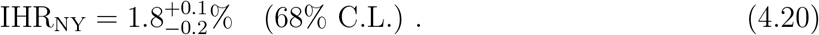

We now turn to the estimation of IFR. While the case fatality rate strongly depends on the number of tests performed, a much more test-independent and intrinsic parameter of COVID-19 is its IFR. Despite its importance, it is difficult to infer the IFR directly from the test results. Various studies (e.g. [4–7]) have estimated its value, which ranges from 0.25% to 1.3% with different methods. Being able to extract the information on the actual number of infections, our methodology and analyses can give an independent and stringent estimate of IFR. Because we have normalized *H*_*N*_ using the hospitalization fatality rate, the value of IFR computed from the NY state is the same for all states. The IFR is therefore the IHR_NY_ multiplied by the hospitalization fatality rate of NY, 0.268, given in Table 4 in Appendix A. From Eq. (4.19), we get the following lower bound on the value of IFR,

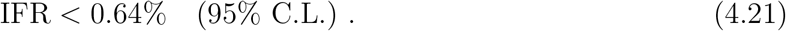

Notice that, although based on the same data set, we are able to obtain a bound on the IFR that is much lower than the overall national average *∼* 5.5%. The overall national average is high because, during most of the state-days, the test volume is relatively low and the IFR is dominated by these data. On the other hand, the data-organizing method used in this paper allows us to systematically map out the parameter space where the most abundant of tests lie, hence derive the information that is blind to the method of simple averaging.

The asymptotic limit of *p*_+_*/H*_*N*_, after extending the theoretical model Eq. (3.14) above the region where the data is available, can give a theoretical estimate for the mean value of IFR,

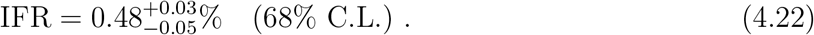

The results (4.21) and (4.22) can be broken down to age-dependent IFRs using the age-dependent statistics provided by [10]. For each age group,

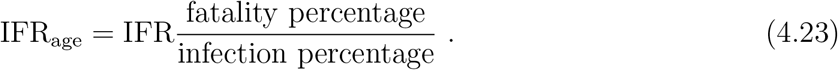

The results are presented in Table 2. The two kinds of percentages used in this calculation are computed in Appendix E.^3^

**Table 2:** Upper bounds (95% C.L.) on IFR and estimates of the values of IFR by age group, followed from the main results Eq. (4.21) and Eq. (4.22).

| age group | $\leq 9$ | 10-19 | 20-29 | 30-39 | 40-49 | 50-59 | 60-69 | 70-79 | 80+ | overall |
| --- | --- | --- | --- | --- | --- | --- | --- | --- | --- | --- |
| IFR (bound) | $< 0.008\%$ | $< 0.008\%$ | $< 0.018\%$ | $< 0.047\%$ | $< 0.11\%$ | $< 0.28\%$ | $< 0.79\%$ | $< 2.0\%$ | $< 3.4\%$ | $< 0.64\%$ |
| IFR (est.) | 0.006% | 0.006% | 0.013% | 0.035% | 0.083% | 0.21% | 0.59% | 1.5% | 2.5% | 0.48% |

We make a comment on an issue related to the symptomatic and asymptomatic cases of COVID-19. In the data we use, the positive cases of viral tests in the USA should so far mostly come from the symptomatic patients. They should also include some asymptomatic cases. The conservative bounds derived above are based on these results. On the other hand, the theoretical extrapolation takes the *T/H*_*N*_ *→ ∞* limit, and therefore should include all symptomatic and asymptomatic infections. In this sense, this is a somewhat bold extrapolation, but can be tested or modified by future data when the test volume gets increased significantly, using the same methodology proposed in this paper.

### 4.3 Deriving the number of hospitalizations from daily tests

As another illustration of the simple relation we obtained, in this subsection, we show how it can be used to derive the number of hospitalizations from the results of the daily viral tests. We also demonstrate under what circumstances this derivation may be more reliable applied.

Knowing the total number of tests (*T*) and the number of positives (*p*_+_), we can plug them into Eq. (3.14), or more generally Eq. (2.13), and solve for *H*. We emphasize here that this equation has multiple roots. What is relevant here is the largest positive root, i.e. the root in the branch that is continuously connected to the point *T/H*_*N*_ = *p*_+_*/H*_*N*_ = 0.^4^

Such predictions are only expected to be rough estimates in terms of order-of-magnitude. Since the relation is derived by combining data from different states, even the order-of-magnitude estimate is expected to be more accurate if used as a national average, or used for states that follow the average behavior of the country relatively well, such as TN and FL. It is less accurate for states that do not follow the average, such as MA and NY. (See Figure 8 for behaviors of the analyzed states.)

To illustrate the above statements with a few examples, we use the new data from Jun 11 to 17 to compare with our predictions from the bestfit model (3.14) fitted with the data collected till Jun 10. In these seven days, in TN state, the total number of tests is *T* = 115746, the number of positive cases is *p*_+_ = 4082. Using the aforementioned method and taking into account of the normalization factor in Table 4, the model predicts a total of *H* = 105 in these seven days. This is to be compared with the recorded *H* = 190 in the database. In the same seven days, in FL state, *T* = 206736, *p*_+_ = 15348, so the model predicts *H* = 510, compared with the recorded *H* = 1052. For AZ state, *T* = 66506, *p*_+_ = 11072, the model gives a bound *H >* 450 (see Footnote 4), compared with the recorded *H* = 385. In these three examples, we see that the model predictions are correct in terms of order of magnitude.

Within the same dates, in MA state, *T* = 59956, *p*_+_ = 1995, the model predicts *H* = 16, compared with the recorded *H* = 343. The prediction is one order of magnitude lower. In NY state, *T* = 442953, *p*_+_ = 4986, the model predicts *H* = 93. Although no new hospitalization data is updated since Jun 3 in the database [2], this prediction is very likely low by an order of magnitude.

We can also apply the model to states that do not have hospitalization data in the database. Again, from Jun 11 to Jun 17, for CA state, *T* = 457190, *p*_+_ = 20824, the model predicts the total normalized-hospitalizations in these seven days to be *H*_*N*_ = 473; for TX state, *T* = 208551, *p*_+_ = 16578, the model predicts *H*_*N*_ = 561 in seven days. For the whole USA, on the day of Jun 17, *T* = 488570, *p*_+_ = 23885, the model predicts *H*_*N*_ = 556 per day.

### 4.4 Projecting actual number of infections

The constant number (*p*_+_*/H*_*N*_)_limit_ that we inferred from the data analyses also provides a simple way to project the total actual number of new infections directly from the number of new hospitalizations each day, without knowing the viral test results. Namely, we have

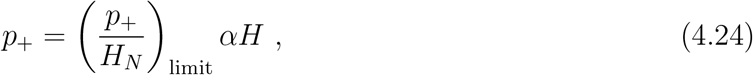

where *α* is the normalization factor of a state defined in Table 4 and *H* is the daily number of the newly hospitalized for the state under consideration.

As we mentioned, although the daily numbers of positive cases are often used as a direct probe of the actual numbers of infections, they are biased due to the test volume which is often not only insufficient but also changing daily. We can use Eq.(4.24) to calibrate these numbers. In Figure 2, we give two examples. In these histograms, we show the daily evolution of the projected actual number of infections versus the confirmed cases. As we can see, besides the differences in the magnitudes, there are also significant differences in the time-dependence. For more examples, see Figure 9.

**Figure 2:**
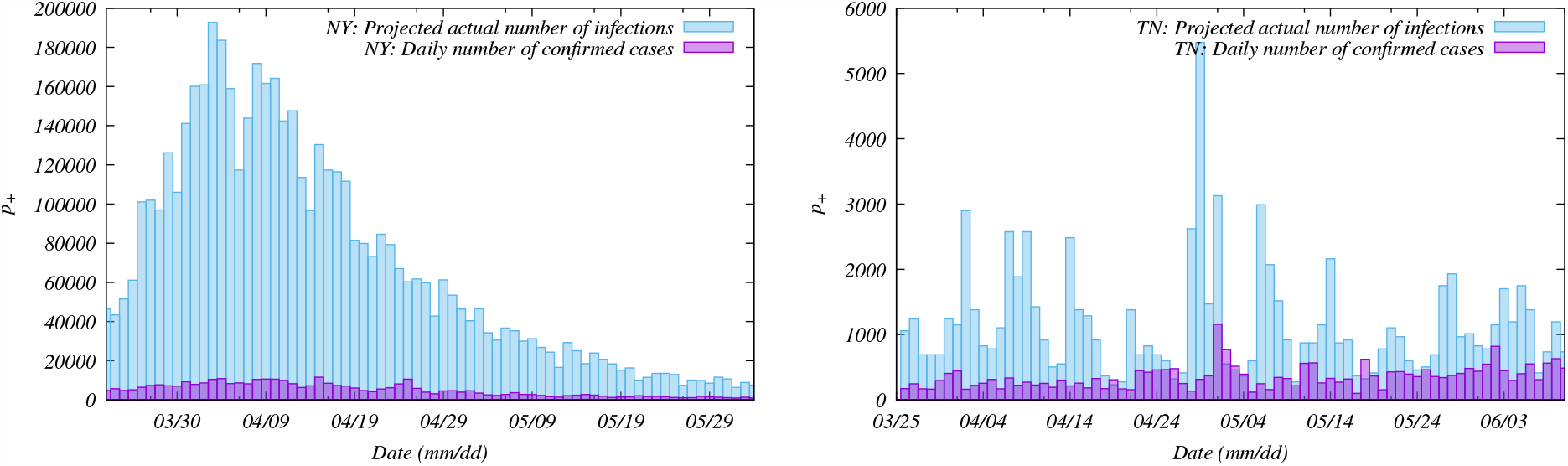
Projected actual numbers of infections versus numbers of confirmed cases in the state of New York (left) and Tennessee (right).

The total actual number of infections can be obtained by summing over the daily values in the histograms, or more simply, directly using (4.24) with the accumulated hospitalization number. The results of several states are summarized in Table 3, in comparison with the numbers of confirmed cases.

**Table 3:** Projected actual total number of infections across different states. The 2nd row presents the total number of hospitalizations, the 3rd row presents the total number of confirmed cases and the 4th row projects the total actual number of infections, till Jun 10, 2020.

| state | NY | FL | MA | GA | TN | OH | VA | MD | WI | MN | AZ |
| --- | --- | --- | --- | --- | --- | --- | --- | --- | --- | --- | --- |
| hospitalization | 89,995 | 11,621 | 10,582 | 8,974 | 1,990 | 6,693 | 7,981 | 9,755 | 2,943 | 3,482 | 3,476 |
| confirmed cases | 374,085 | 67,371 | 104,156 | 53,980 | 28,061 | 39,575 | 52,177 | 59,465 | 21,593 | 30,199 | 29,852 |
| projected infections | 5,070K | 608K | 1,570K | 490K | 92K | 516K | 319K | 599K | 141K | 267K | 231K |

**Table 4:** Hospitalization Fatality Rates (HFR) and normalization factors *α* of different states. The second row is the total number of hospitalizations, the third row is the total number of fatalities, till Jun 10, 2020 [2]. (NY is up to Jun 3, because there is no update on the numbers of hospitalization after that [2].) The fourth row is the values of HFR. The last row is the normalization factor, *α*, used to normalize the value of the daily numbers of hospitalizations, *H*, using HFR. After the normalization, *H*_*N*_ = *αH*.

| state | NY | FL | MA | GA | TN | OH | VA | MD | WI | MN | AZ |
| --- | --- | --- | --- | --- | --- | --- | --- | --- | --- | --- | --- |
| hospitalization | 89,995 | 11,621 | 10,582 | 8,974 | 1,990 | 6,693 | 7,981 | 9,755 | 2,943 | 3,482 | 3,476 |
| fatality | 24,079 | 2,889 | 7,454 | 2,329 | 436 | 2,457 | 1,514 | 2,844 | 671 | 1,267 | 1,095 |
| $\frac{\text{fatality}}{\text{hospitalization}}$ | 0.268 | 0.249 | 0.704 | 0.260 | 0.219 | 0.367 | 0.190 | 0.292 | 0.228 | 0.364 | 0.315 |
| $\alpha$ | 1 | 0.929 | 2.63 | 0.970 | 0.817 | 1.37 | 0.709 | 1.09 | 0.851 | 1.36 | 1.18 |

The total actual number of infections can also be projected using the accumulated number of fatalities using the estimated IFR value in (4.22), *N* = total fatalities*/*IFR. Although there is a time-delay, this is negligible for estimations of total numbers. This method is particularly useful for states that do not have hospitalization data in the database. For example, till Jun 10, the total fatality in the state of CA is 4776. This projects a total of 995K infections, compared to the confirmed 136K.

With these information, we can also estimate the percentage of the population that have been infected. To give a few examples, recall the current total populations of NY, MA, CA and TN are 19.5M, 6.89M, 39.5M and 6.83M, respectively. With the projected infections computed above, till Jun 10, respectively, approximately 26%, 23%, 2.5% and 1.5% of the population have been infected.

## 5 Discussions

In this paper, we have studied the bias between the number of confirmed cases and actual number of infections of COVID-19, focusing on understanding the dependence of this bias on several variables, such as the number of total tests, confirmed cases, hospitalizations and fatalities. Using a simple model, we introduce a strategy to organize the historical data of the USA on these variables. After considering theoretical constraints and fitting the model with the data, we find a simple formula relating the mean values of these variables. We show several applications of this main result, such as the estimations of the actual number of infections and the value of IFR of COVID-19.

In this section, we discuss several issues related to our analyses. In the analyses so far, we assume, for a hospitalized patient, the test result and the hospitalization are reported on the same day on average. However, there might be some delay between these two events. The CDC [8] reports that 50% of people take 3-7 days from symptom onset to seeking out-patient care, and the mean number of days from symptom onset to hospitalization (standard deviation) is around 7 (5) days. So the mean delay, if any, between the reporting of the viral test result and hospitalization should be much fewer than 7 days. To test if there is any significant effect on the main results from this delay, in Appendix F, we shift the relative dates between these two events by 1, 2, 3, 5 days, respectively, and repeat the same analyses. As we can see, the qualitative conclusions remain the same. In particular, the upper bound on IFR is quite robust, while the lower limit of the theoretical prediction of IFR may be lowered. We expect that most of the variations between these trial cases with different delays come from daily fluctuations of the hospitalization numbers. Since we do not expect the average number of hospitalization to change much over several days, a smoothing of the data over a few days, before performing our analyses, might reduce these fluctuations and is an interesting subject for future study. Nonetheless, we point out that, if we compare the daily evolution of *H*(*t*) and *p*_+_(*t*) in Figure 2 and Figure 9, even for those states that have clear peaks in the evolution, there is no evidence that there is a time dealy.

Another application is to use the results of the USA obtained in the paper to make predictions for other regions or countries that are still in the middle of the pandemic but without sufficient tests. While there might be differences in the conditions that lead to hospitalization and fatality, some simple modification of our normalization procedure may make the results easily adapted to a different situation. It is also worth to analyse the data of different countries using the same methodology of this paper.

The current model, especially Eq. (3.14), is very simple. With more data, it would be interesting to check its predictivity and to see if it is necessary to make it more sophisticated. Also, since the impact of the disease gets much worse with increasing age, it would be desirable to study the age-dependence of the bias.

## Data Availability

All data come from https://covidtracking.com/

## Acknowledgments

We would like to thank the volunteers of the COVID Tracking Project [2] and the COVID project of 1Point3Acres.com [3] for their vision and devotion in creating these database and for collecting daily the valuable data from many resources. We thank Alyssa Goodman, Avi Loeb, Jiong Lu, Dongliang Qian, Yun Zhang, Qiang Zhou, and Wenzhan Zhou for helpful discussions.

## A Hospitalization fatality rate and normalization of *H*

The hospitalization fatality rates (HFRs) mentioned in subsection 2.1 are computed in Table 4. Note that, in computing HFR, we have used the most recent ratios of the total number of fatalities to that of hospitalizations. As demonstrated in Figure 3, these ratios have stabilized over a period that is much longer than the typical length of delay between the hospitalization and fatality, and therefore can be used as the time-independent and characteristic parameters of the respective states.

**Figure 3:**
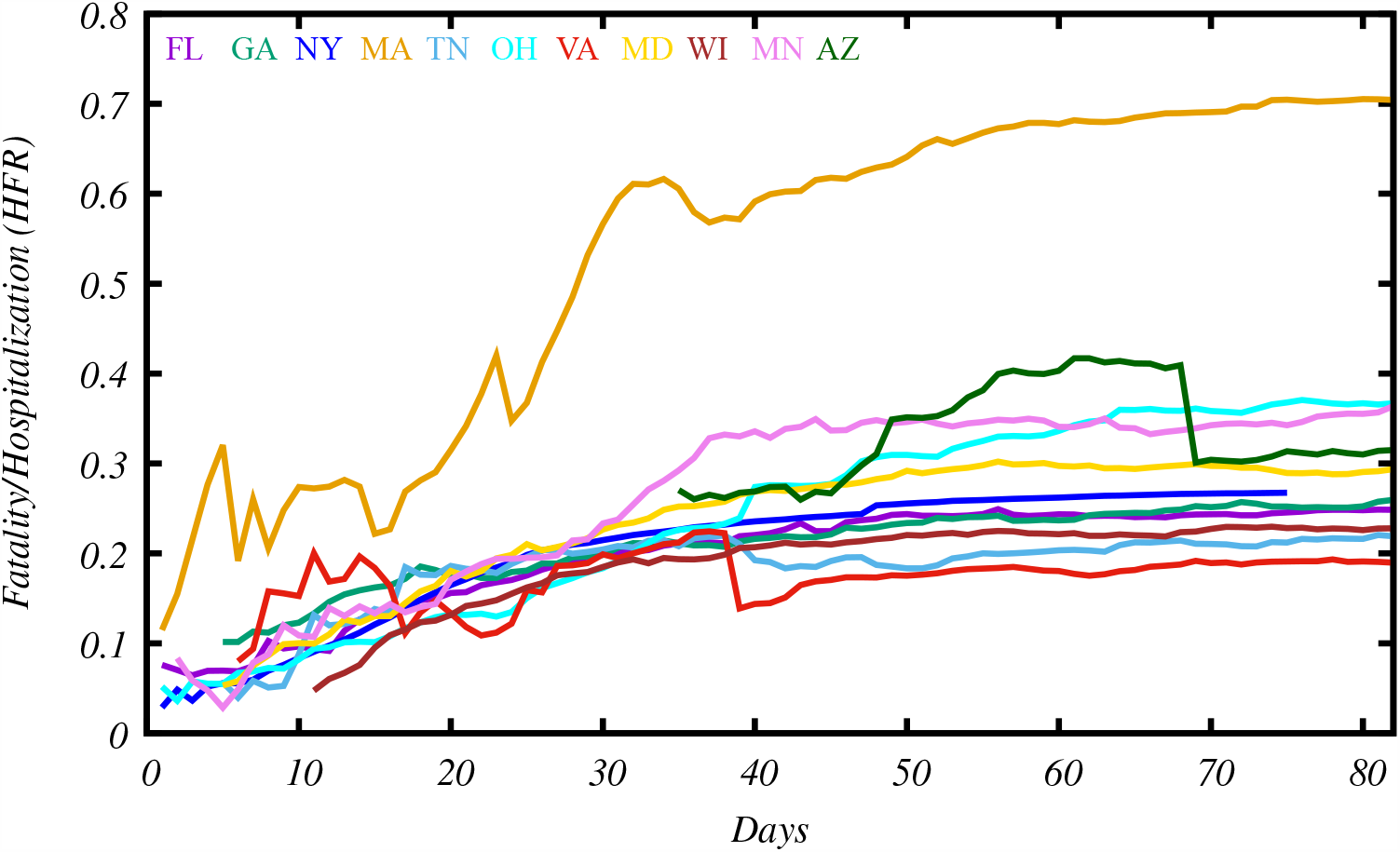
The evolution of HFR as a function of time till Jun 10, 2020 (except for NY, till Jun 3), for different states. Note that, for all states, the changes in the rates freeze after about 70 days since the first reported hospitalization.

As mentioned in subsection 2.1, we use HFRs to normalize the value of *H* to *H*_*N*_,

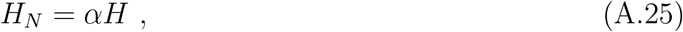

so that, with the normalized values *H*_*N*_, all states would have the same HFR. The normalization constants *α* that satisfy this requirement are given by

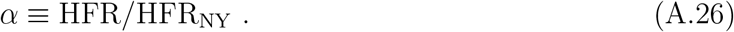

The normalization factors are different across the states, and we use NY state as the reference point so that *α* = 1 for NY. The values of *α* for other states analyzed in this paper are given in Table 4.

## B Data resources

The data we used in this paper are obtained from the historical data collected in the website of the COVID Tracking Project [2]. In the historical data of each state, the numbers of total tests *T* are given by the daily records of “New Tests”. Numbers of confirmed cases are recorded accumulatively in this website, so to get *p*_+_, we need to subtract the value of “Cases” of the day by that of the previous day; and the same for the number of hospitalizations *H* and the number of fatalities *D*, from the column “Hospitalized” and “Deaths”, respectively. To make the subtraction, we start the analyses from the 2nd day on which such data become available.

We have used 789 state-days of data points from 11 states in the USA recorded from the end of March to Jun 30. Among them, there are a small number of glitches, involving zero and negative numbers, that need to be adjusted to be used in our analyses.

- Sometime due to testing or reporting strategy changes, the website reports a negative number on one day, reflecting the over-counting of the previous day(s). In these cases, we combine the values of this day with a neighbouring date. (e.g. Starting from May 2, VA started to list separately the numbers of “total people tested” and “unique people tested” because some people were tested twice. April 18 of MA and May 15 of AZ have negative increases in total Hospitalized.)
- A special case is May 27 of GA, on which day the state started to report the confirmed number of antibody tests included in all past tests, which resulted in a large negative number in the total test, impossible to be combined with any previous dates. We skip this date after taking care of the continuity of the other numbers to the next day. The unspecified error of including the number of antibody tests in all past dates should be relatively small, because the total number of antibody tests is much smaller than that of viral tests. Such errors should eventually lead to more statistical scattering of the data.
- If the tests are 0 on certain dates, we remove those dates.
- If we have N/A hospitalization in the middle of the column, we add the tests to the date where the hospitalization is available (e.g. VA: April 11-15).
- If we have 0 hospitalization in the middle of the column, we add the tests to the date where the hospitalization is available (AL: April 20 and 23)
- MD started to regularly report the negative cases in addition to the positive cases on May 28, so we start to use the data from May 29.
- NY’s data is used up to Jun 3, because the hospitalization data stops being updated after that.

Besides these, there are still another small number of glitches, which might be due to human errors. For example, in some dates, some state website might have listed the number of new cases, but forgot to update the total number of tests; and the volunteers of the COVID Tracking Project chose to assume the latter to be the same as the former. In some other cases, there are very large fluctuations in the number of tests which might be due to under-reports in certain dates and re-grouping in other dates. We do not make any modifications in these situations, as long as they are all positive numbers. These possible errors may contribute to part of the statistical errors in our analyses.

## C Some data analyses details and plots

In this appendix, we supply some details of data analysis methods and some intermediate results used in the main text of the paper.

From 11 states we gather data for 805 state-days after applying our selection criteria described in Appendix B. Note that here for each day we make use of *T, p*_+_, and *H* and therefore 2415 data points. At the same time, we normalize all values of *H* according to the procedure described in Appendix A. After normalization, we bin *T/H*_*N*_ data in different *p*_+_*/H*_*N*_ bands.

Due to the sparse nature of the data, we use variable bin-widths at different ranges of *p*_+_*/H*_*N*_. We change the bin-widths at *p*_+_*/H*_*N*_ = 20 and 50. The data points are binned with a width of Δ*p*_+_*/H*_*N*_ = 2 till *p*_+_*/H*_*N*_ = 20. After *p*_+_*/H*_*N*_ = 20 we use a band of Δ*p*_+_*/H*_*N*_ = 5. Note that there are only 16 data points beyond *p*_+_*/H*_*N*_ = 50 and therefore beyond that point we make groups of 5 data points together regardless of bin-width.

We provide a histogram of *T/H*_*N*_ within three initial bands of *p*_+_*/H*_*N*_ in the top left plot of Figure 4. In this figure, we use a binwidth of 5 to show more data points. This hisogram importantly reveals that the distribution of *T/H*_*N*_ is significantly skewed. In order to search for a basis where the distribution is normal, we explore three scaling strategies, vis.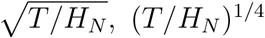 and log(*T/H*_*N*_). The corresponding histograms are plotted in top right, bottom left and right respectively. Note that while the power law scaling helps to reduce the skewness, in logarithmic scaling of *T/H*_*N*_, we obtain the histogram close to normal distribution in all 3 bins within *p*_+_*/H*_*N*_. We obtain the mean of the mean 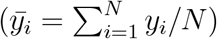 and 1*σ* error of the mean 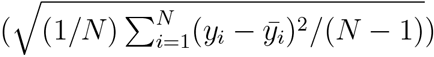, where *y*_*i*_ = log(*T/H*_*N*_). We then convert the mean values of *y*_*i*_ and the errors of the means to those of *T/H*_*N*_. Using the obtained values, we plot the binned data with 1*σ* error of the mean in Figure 5. In the top panel we plot the data for all states in different colors with the binned data for the entire range. Note that beyond *p*_+_*/H*_*N*_ = 50, we identify only 16 scattered data points within *p*_+_*/H*_*N*_ = 50 *−* 200. At the same time, contrary to the theoretical constraint stated below Eq. (2.13), the data points when binned, show a decreasing trend when *T/H*_*N*_ increases. These points indicate very low hospitalization despite large positive cases obtained with relatively small number of testing. Considering the various possibilities of systematic errors mentioned in Appendix B, we do not include the data points beyond *p*_+_*/H*_*N*_ = 50 in our analyses. With more data at the low positive rate region it will be possible to perform better statistical analysis. In the bottom panel of the same figure we plot an enlarged version of the top panel where we only highlight the part used for our analysis. In this plot, a non-linear increase in *T/H*_*N*_ is evident as *p*_+_*/H*_*N*_ increases, as expected from the theoretical constraint that *p*_+_*/H*_*N*_ should approach an asymptotic limit.

**Figure 4:**
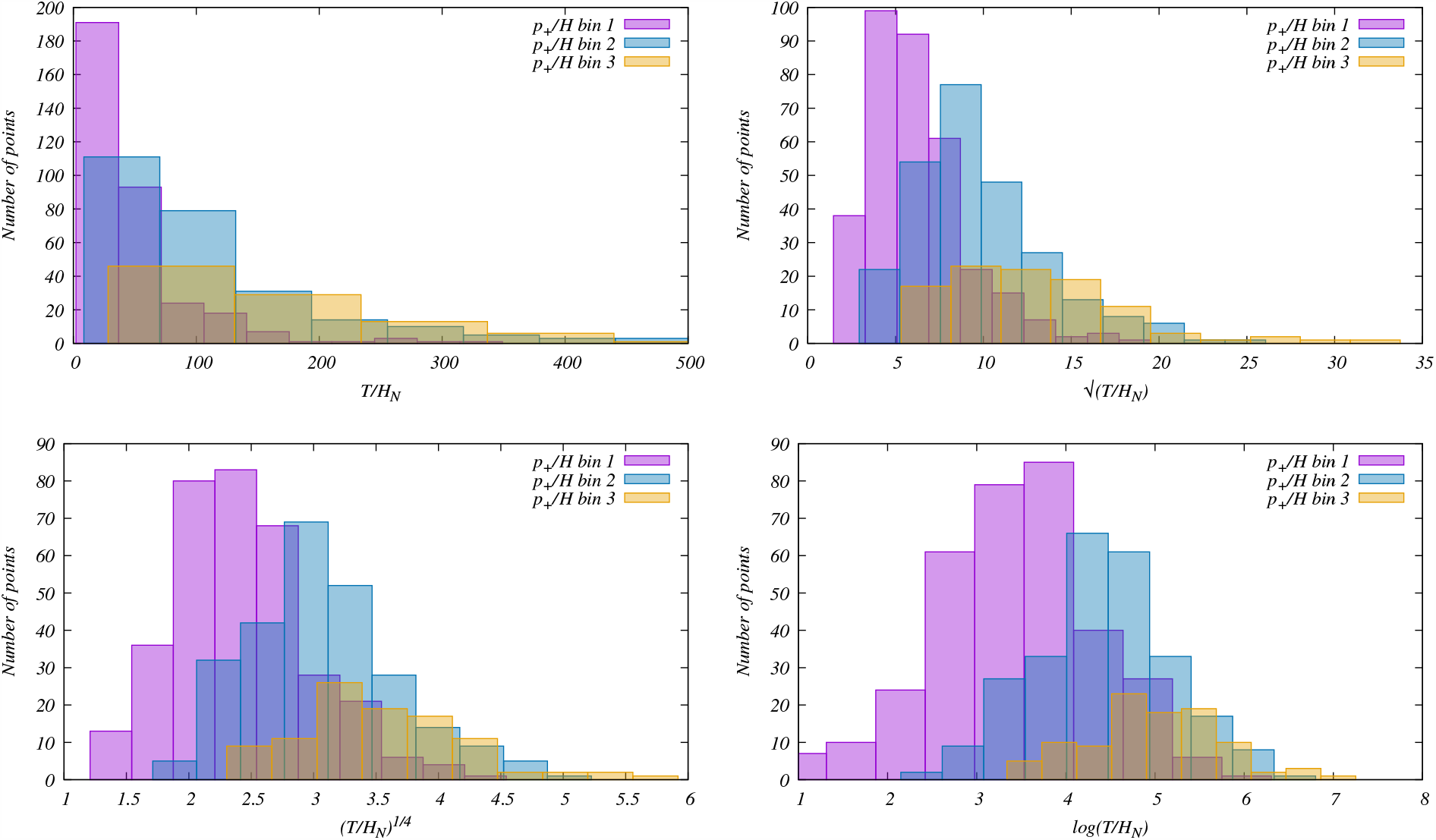
We provide the distribution of *T/H*_*N*_ data in first three *p*_+_*/H*_*N*_ bins with a binwidth of 5. Histograms of *T/H*_*N*_ (top left), 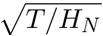 (top right), (*T/H*_*N*_)^1*/*4^ (bottom left) and log(*T/H*_*N*_) (bottom right) are provided. Note that while distributions of *T/H*_*N*_ in these three bins are significantly skewed, rescaling helps to reduce the skewness and the distribution of log(*T/H*_*N*_) is closest to normal.

**Figure 5:**
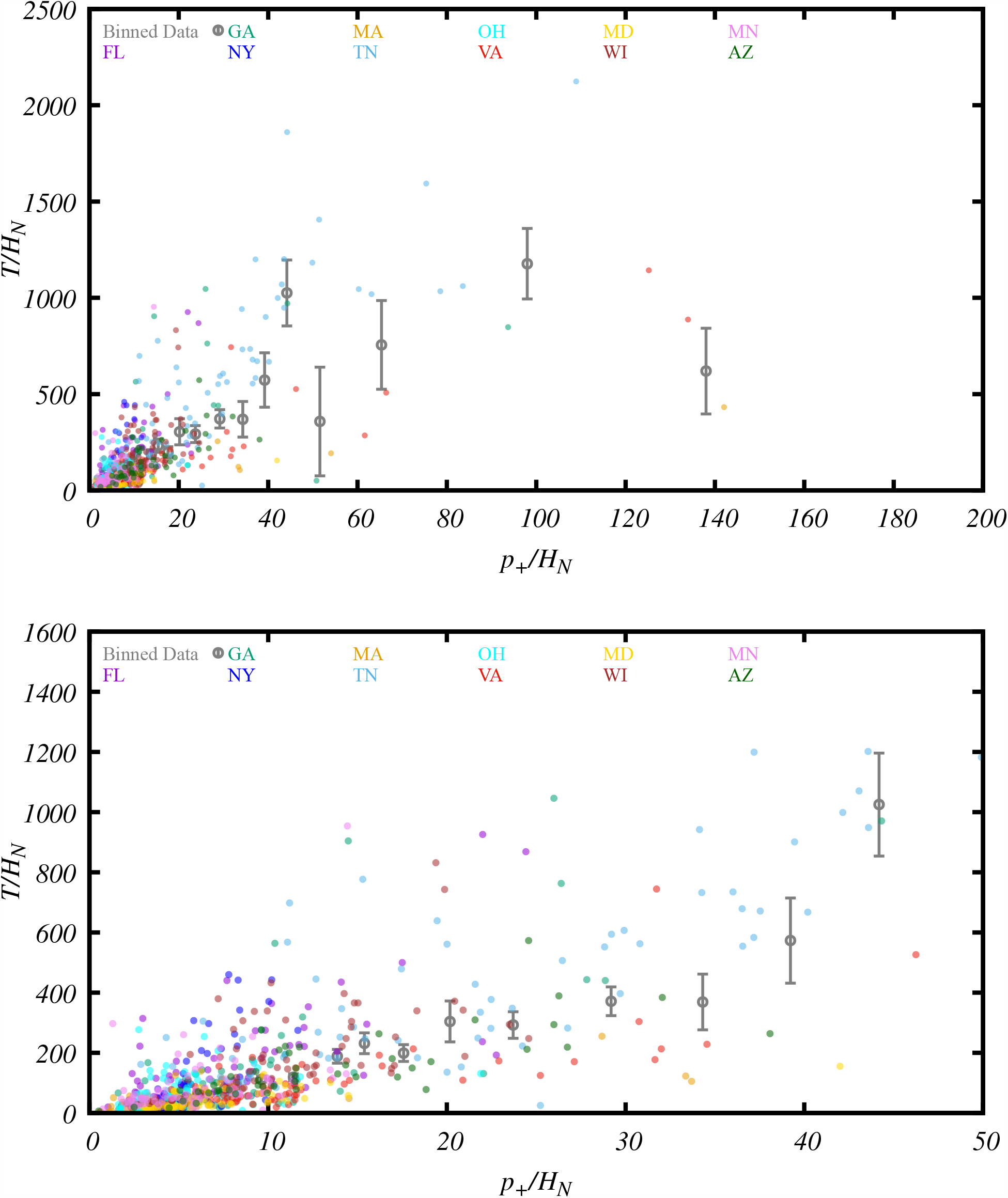
Unbinned data points in different colors are plotted from 11 states for 805 state days (top panel). Binned data points are obtained with different binwidths at different region of *p*_+_*/H*_*N*_ as mentioned in the text. A zoomed version of the top panel is provided at the bottom panel that represents the dataset used in this paper.

**Figure 6:**
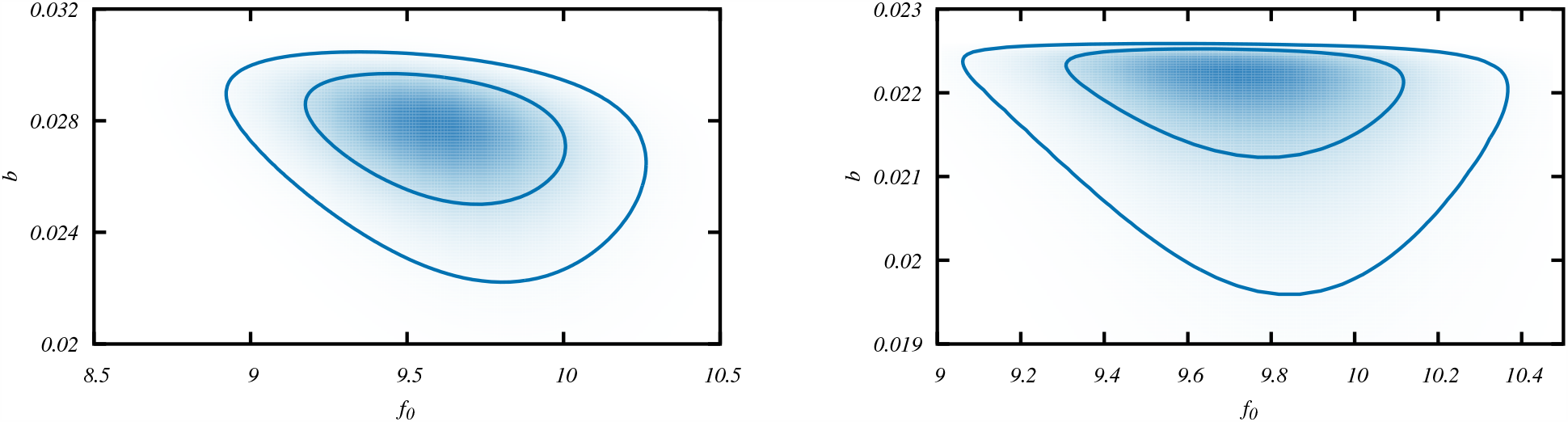
Correlation between parameters *f*_0_ *− b* when the model Eq. (3.14) (left) and Eq. (C.27) (right) are used to compare with the data. Note that the contour for the ArcTanh function gets sharply cut off because this function approaches the limit exponentially fast.

Therefore with 789 state-days, we perform our fit to the binned data. Here we do not include correlation between different bins. We use NLOPT [9] to obtain the best fit functions by minizing the *χ*^2^. We presented the result for the tangent model (3.14) in Sec. 3. For the inverse hyperbolic tangent function,

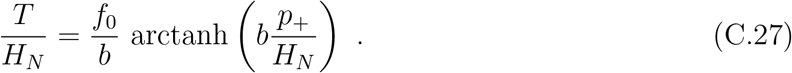

the best-fit parameters are *f*_0_ = 9.71, *b* = 0.022. Similarly, if we assume that the fitting function is only valid up to the last binned data point, according to this model,

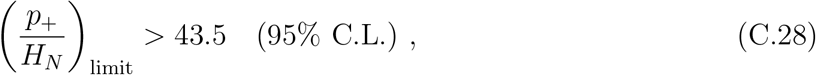

If we take the theoretical extrapolation of this model, we can get the asymptotic value of *p*_+_*/H*_*N*_ as

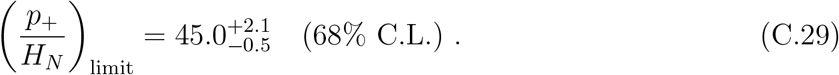

The correlation between the parameters *f*_0_ *− b* in different cases are obtained with grid search in conservative prior ranges in both the parameters. We use the *χ*^2^ differences criterion to find the 68% and 95% confidence regions for 2 parameters. We plot the 2D likelihood marking 68% and 95% regions in Fig. 6. Results for Eq. (3.14) and Eq. (C.27) are plotted at the left and right panel, respectively. Note that compared to Eq. (3.14), the probability of parameter *b* in Eq. (C.27) sharply drops at higher *b* region because of the exponential behavior in the ArcTanh function. From the samples within 68% and 95% regions we identify the error-bands on *T/H*_*N*_ as a function of *p*_+_*/H*_*N*_ that are plotted in Fig. 1 and in Fig. 7. As expected, the lower error-bands for ArcTanh (corresponding to large values of *b*) case closely overlap as the distribution falls rapidly. Note that while from the 2D likelihood we obtain bounds for both parameters *f*_0_ and *b*, only the errors on *b* determine those of the asymptomatic limits of *p*_+_*/H*_*N*_.

**Figure 7:**
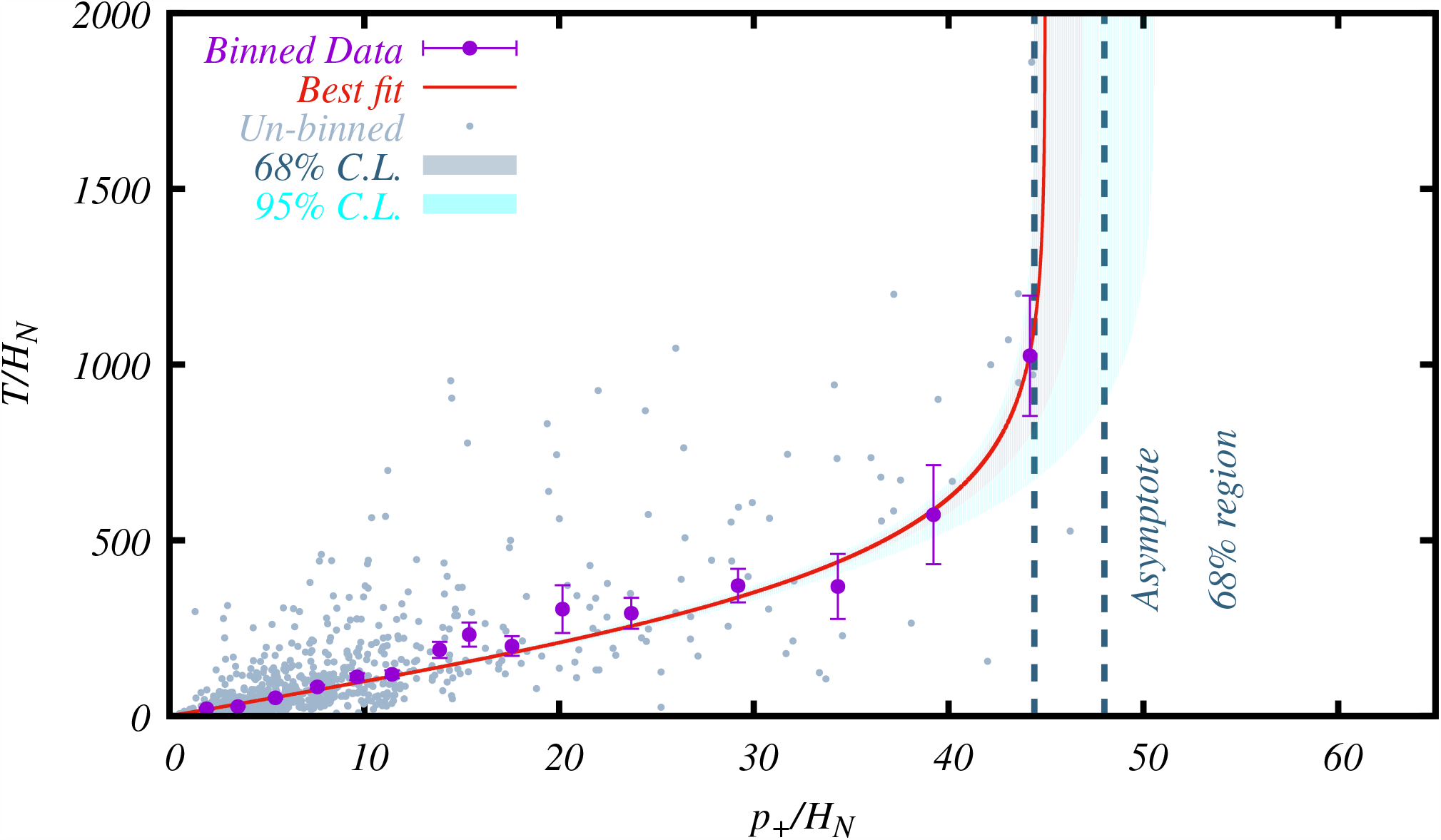
Same as Figure 1 but fitted with the inverse hyperbolic tangent function Eq. (C.27).

**Figure 8:**
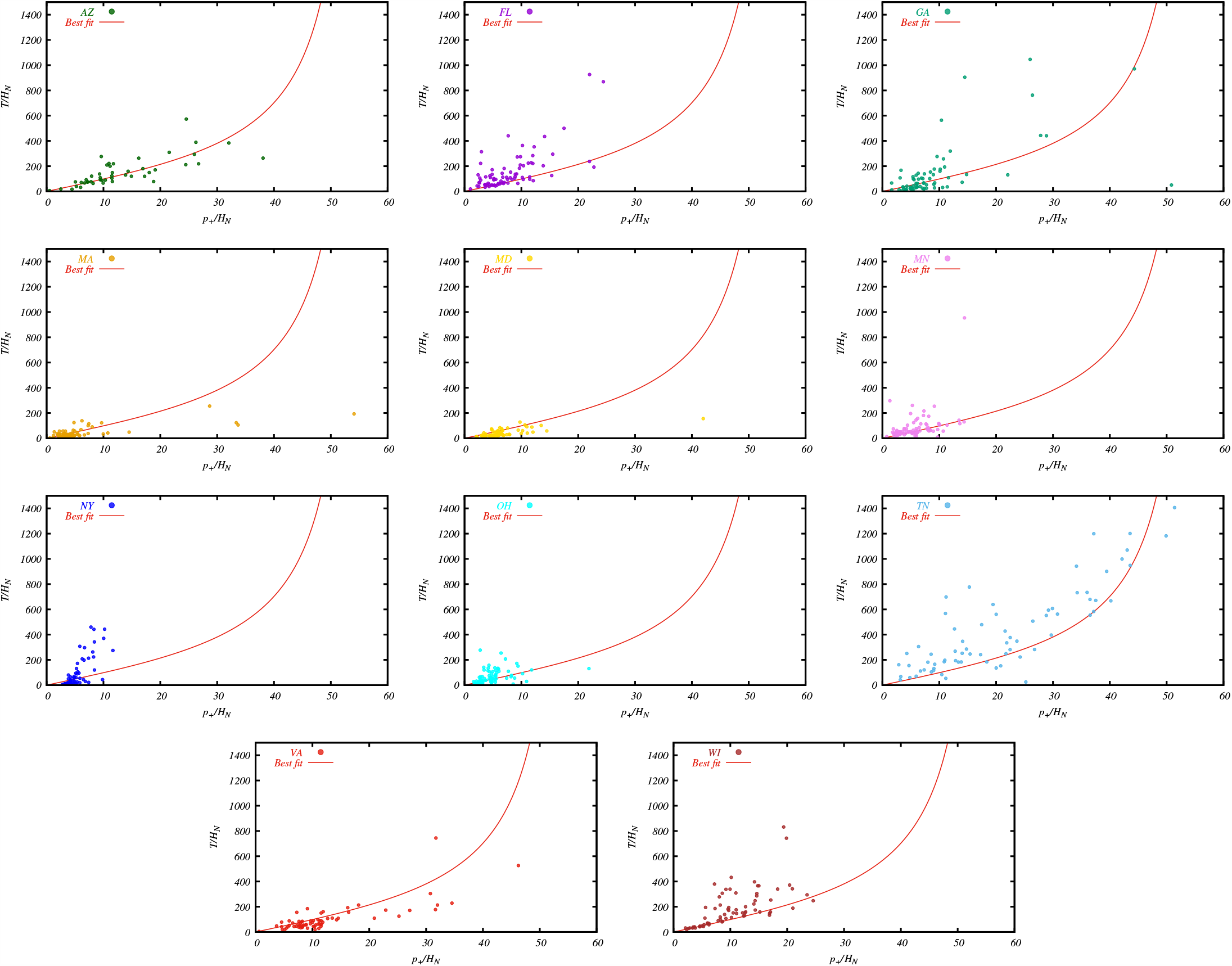
Unbinned data from each state against the best-fit curve to the mean.

**Figure 9:**
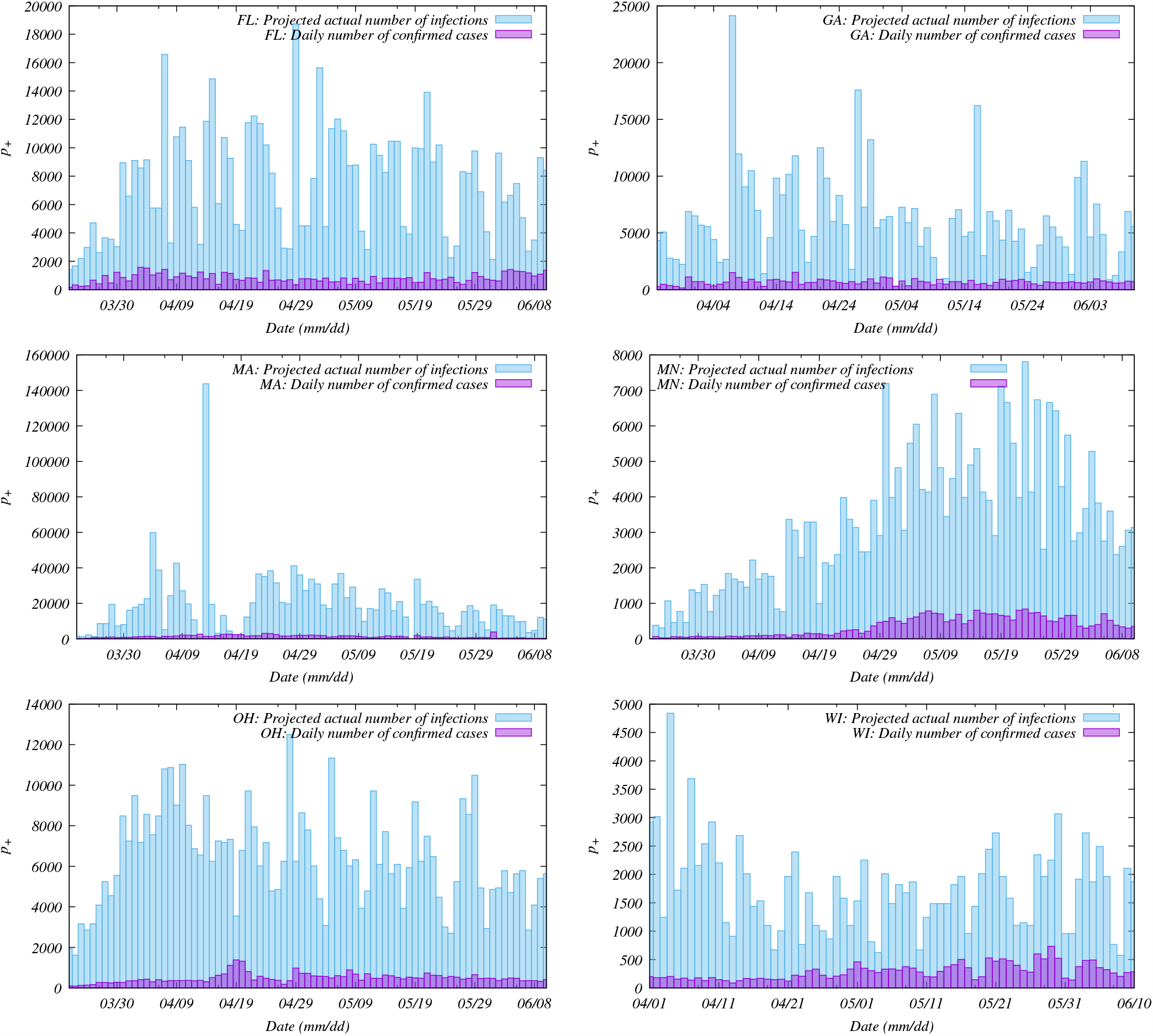
Daily values of the number of confirmed cases versus the actual number of infections projected from Eq. (4.24), for the state of Florida (top left), Georgia (top right), Massachusetts (middle left), Minnesota (middle right), Ohio (bottom left), and Wisconsin (bottom right).

The *χ*^2^ of the ArcTanh model is higher than that of the Tan model by 1.2, so the two models are statistically indistinguishable by the current data. More data with small positive rates could help to distinguish the two models in the future. These two models predict slightly different limiting value for *p*_+_*/H* and the ArcTanh model gives a slightly higher value of IFR, although they are statistically consistent within 2*σ* errors. These two functions also approach the limiting value of *p*_+_*/H*_*N*_ with different speeds – in power-law for the model (3.14) and exponentially fast for the model (C.27).

## D Some plots about individual states

In this appendix, we supply some extra plots helpful to the discussions in the main text. In Figure 8, we plot the unbinned data of each of the 11 states used in this analyses against the best-fit curve to the mean. These plots help to illustrate the differences between the individual states and how they are distributed around the best-fit.

In Figure 9, we show, for some selected states in addition to Figure 2, the daily evolution of the number of the confirmed cases versus the actual numbers of infections projected from Eq. (4.24). Since the daily evolution of number of the confirmed cases is *p*_+_(*t*) and the projected actual number of infections follows from *H*(*t*), these histograms can also be used to examine if there is any systematic time delay between the posting of the viral test result and the hospitalization event for the same patient. (However, note that here *p*_+_(*t*) includes all confirmed cases among which many are not hospitalized.) On average, there is no obvious delay observed.

## E Infection and fatality percentage by age group

In this Appendix, we list in Table 5 the statistics about the case percentages and fatality percentages by age group, which are used in the computation of the IFRs by age group in subsection 4.2. Note that, in the age group calculation in subsection 4.2, what is needed is the infection percentages by age group, we assume they are the same as the case percentages for all age groups. This assumption assumes that the ratio of the number of confirmed cases to the actual number of infections, i.e. the bias, is the age-independent. If the bias is age-dependent, e.g., lower in younger people and higher in older people, the IFRs by age group computed in subsection 4.2 would become even smaller for younger people and higher for older people. However, we currently cannot find the data that can be used to analyze this age-dependent property.

**Table 5:** Numbers of confirmed cases and fatalities by age group in the USA till May 30, 2020, from [10], and the resulted percentages by age group.

| age group | $\leq 9$ | 10-19 | 20-29 | 30-39 | 40-49 | 50-59 | 60-69 | 70-79 | 80+ |
| --- | --- | --- | --- | --- | --- | --- | --- | --- | --- |
| number of cases | 20458 | 49245 | 182469 | 214849 | 219139 | 235774 | 179007 | 105252 | 114295 |
| case percentage | 1.5% | 3.7% | 13.8% | 16.3% | 16.6% | 17.9% | 13.6% | 8.0% | 8.7% |
| number of fatalities | 13 | 33 | 273 | 852 | 2083 | 5639 | 11947 | 17510 | 32766 |
| fatality percentage | 0.018% | 0.046% | 0.38% | 1.2% | 2.9% | 7.9% | 16.8% | 24.6% | 46.1% |

## F Time delay between case confirmation and hospitalization

In our main analyses, for each unbinned data point in the plots, we use the data of *p*_+_(*t*_1_), *T* (*t*_1_) and *H*(*t*_2_) on the same date, *t*_1_ = *t*_2_. Following the discussion in Sec. 5, if on average there is a time delay between the posting of the viral test result and the hospitalization for the same patient, then we should use *p*_+_(*t*_1_), *T* (*t*_1_) and *H*(*t*_2_) with different dates instead. In this appendix, we test if this change would significantly affect our conclusions.

While in usual practice, positive cases with severe symptoms are hospitalized on the day of detection, there might be certain time lag between sample collection, test and hospitalization. Assuming such a delay exists and its average value is *d* days, we apply the same delay to all the grouping of *p*_+_(*t*_1_), *T* (*t*_1_) and *H*(*t*_2_) that are used to generate the unbinned data point. Namely, we assume that the data of hospitalizations, *H*(*t*_2_), corresponds to the cases detected *d* days ago, so the data about *T* (*t*_1_) and *p*_+_(*t*_1_), which are used together with *H*(*t*_2_) to generate the unbinned data point, should have the time coordinate *t*_1_ = *t*_2_ *− d*. After this time shift, the procedure of the rest of the analyses is the same as what is described in Appendix C.

We test four different trials, where *d* = 1, 2, 3, 5 days. The results are presented in Figure 10, 11, 12, and 13, respectively. We present all unbinned and binned data points in the top panels of these figures. Similar to the no-delay case, if *p*_+_*/H*_*N*_ *>* 50, we have several scattered binned data coming from significantly small number of unbinned data points over large bin sizes, and their behavior do not satisfy the theoretical constraint stated below Eq. (2.13). Considering systematic errors mentioned in Appendix B, in the bottom panels, we cut out these binned data points and present the best-fit curves of the model (3.14) to all binned data points below *p*_+_*/H*_*N*_ *<* 50. In the captions, we also give the 68% ranges of the asymptotic limits of *p*_+_*/H*_*N*_. We find that, for the best fit, *χ*^2^ = 25 *−* 35 for 15-16 binned data points in all cases, including the no-delay case. We find that the lower bounds on (*p*_+_*/H*_*N*_)_limit_ based on data fitting in all the cases are consistent with the no-delay case, Eq. (3.16). We also find that the linear growth behavior in the function *F* (*p*_+_*/H*_*N*_) is not allowed in any of the cases at 95% C.L.. However, we find that, for the trials of 1, 2 and 3 days of delay, based on theoretical extrapolation, the upper 68% limits for the value of (*p*_+_*/H*_*N*_)_limit_ are higher, although the lower 68% limits are similar. For example, the upper 68% limit for (*p*_+_*/H*_*N*_)_limit_ can be as high as 106, which implies that the lower 68% limit of IFR is as low as 0.25%. The 5 day delay trial is found to be very similar to the no-delay case. However, it should be noted that 5 days delay in hospitalization of severe cases after detection may not represent a realistic scenario on average.

**Figure 10:**
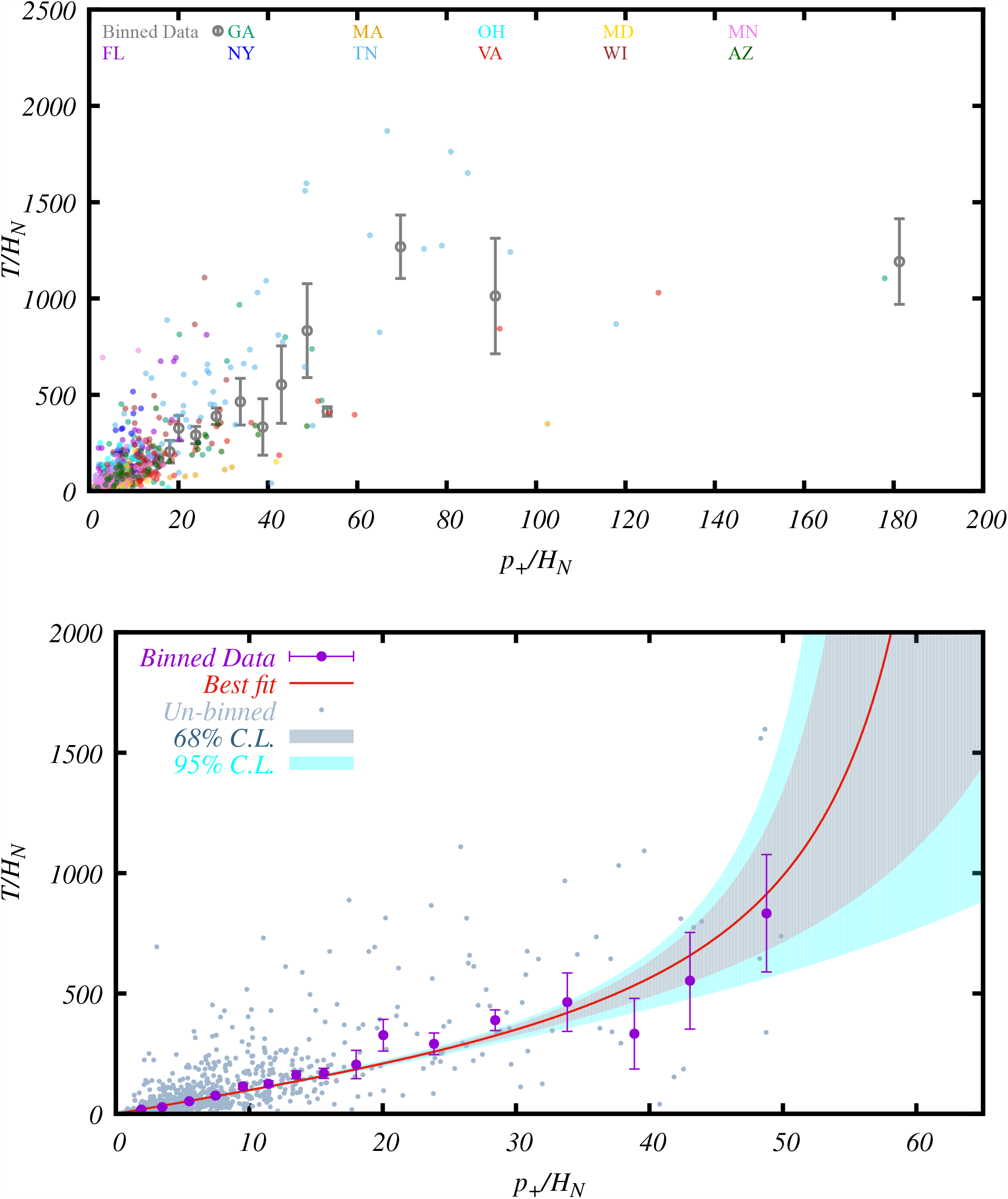
The same as Figure 5 but assuming that there is on average one day delay between the posting of the test result and the hospitalization. In the bottom panel, the cut is applied and the best fit model of Eq. (3.14) is plotted. The 68% C.L. region for (*p*_+_*/H*_*N*_)_limit_ is between 60 *−* 83.

**Figure 11:**
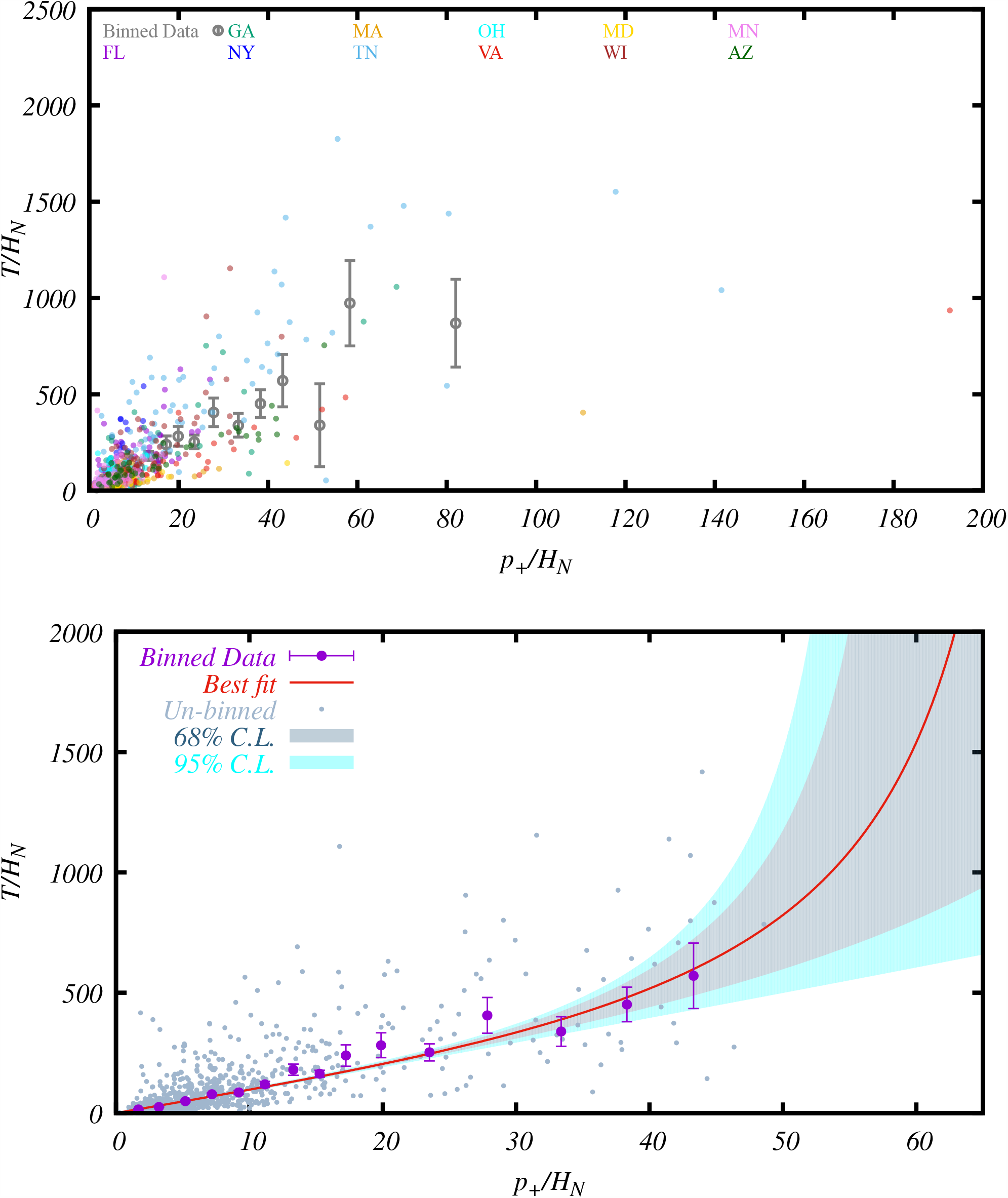
The same as Figure 10 but assuming two days of delay. The 68% C.L. region for (*p*_+_*/H*_*N*_)_limit_ is between 62 *−* 106.

**Figure 12:**
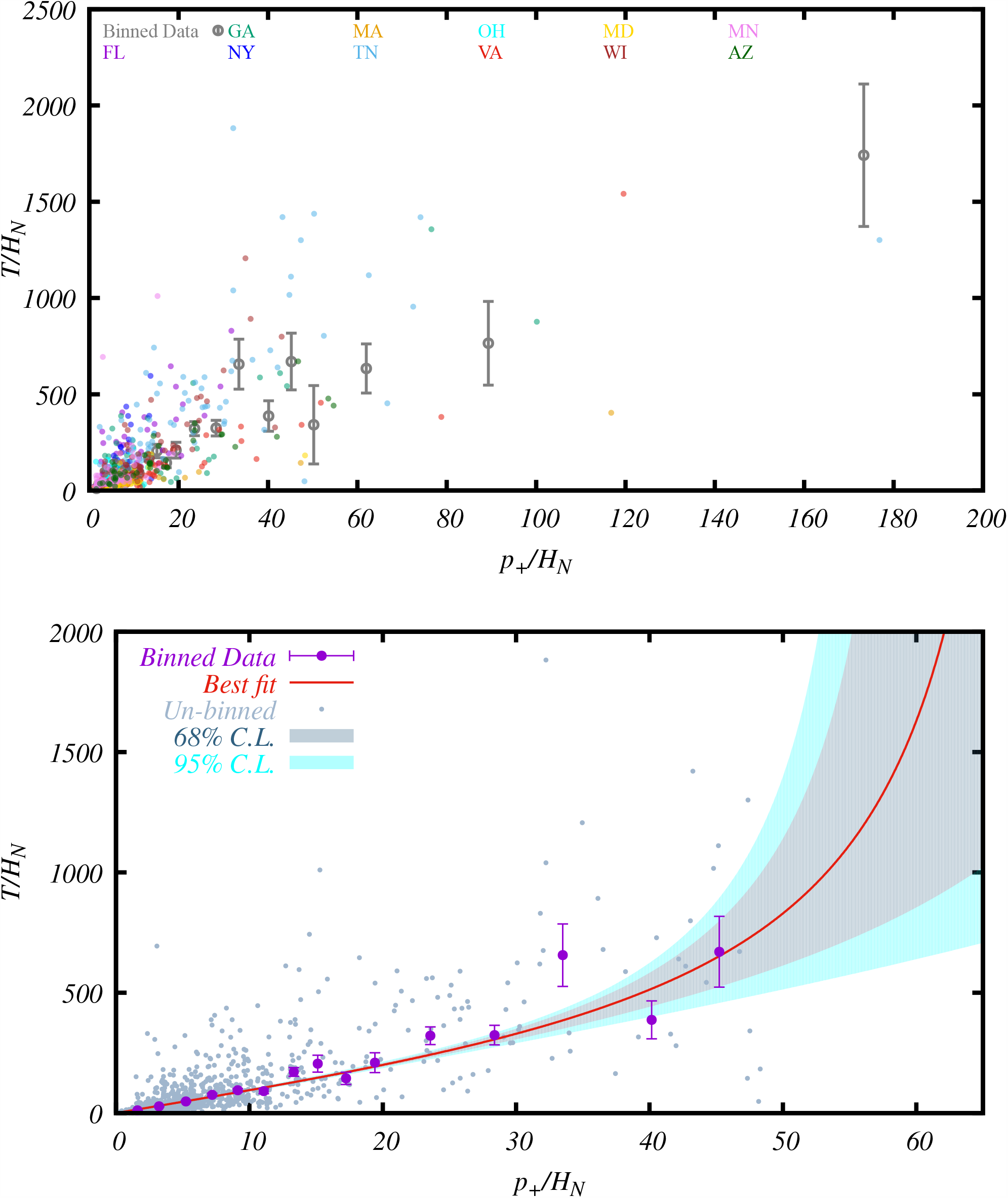
The same as Figure 10 but assuming three days of delay. The 68% C.L. region for (*p*_+_*/H*_*N*_)_limit_ is between 62 *−* 97.

**Figure 13:**
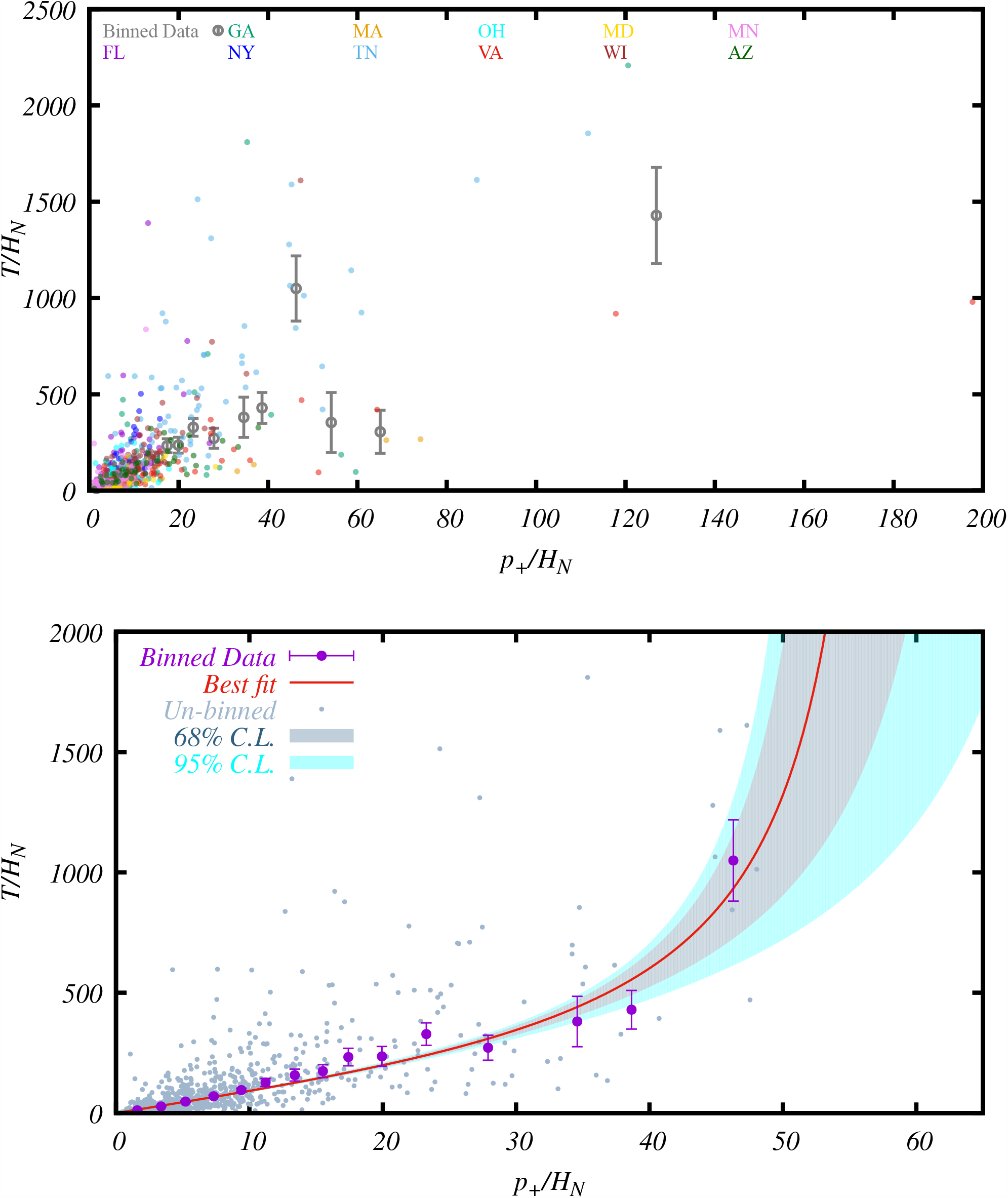
The same as Figure 10 but assuming five days of delay. The 68% C.L. region for (*p*_+_*/H*_*N*_)_limit_ is between 56 *−* 68.

As pointed out in section 5, because we expect the change of the average number of hospitalizations to be small over a few days, the variations between the above different trials may be due to statistical fluctuations, and a smoothing treatment of the raw data before the analyses may help.

## Footnotes

1 In this paper we quote 68% confidence level (C.L.) regions on measured parameters and 95% on lower bounds.

2 For the model (3.14), the largest positive rate is 1*/f*_0_ in terms of national average. If *r*_+_ *>* 1*/f*_0_, the root is taken to be zero.

3 When using (4.23), we have assumed that the bias, i.e. the ratio of the number of confirmed cases to the actual number of infections is age-independent, since we do not have data that can be used to analyze the age-dependence in the bias.

4 Because the maximum positive rate of the best-fit is 1*/f*_0_, for a test with *r*_+_ *>* 1*/f*_0_ *≈*0.104, there is no root in the mentioned branch and the roots in other branches should not be used. In this case, we use the values at *r*_+_ = 0.1 to provide a bound for *H*_*N*_. See its value in the first column in Table 1. Namely, *H*_*N*_ *>* 0.1*T/*12.5. For such an example, see the case of AZ state below.

